# Comprehensive proteomic characterization of the esophageal String Test identifies a luminal protein signature associated with eosinophilic esophagitis

**DOI:** 10.64898/2026.09.30.26364161

**Authors:** Flor Delgado-Cruz, Francisca Molina-Jiménez, Sergio Casabona, Elena Resina, Verónica Martín-Domínguez, Jennifer Fernández-Pacheco, Laura Arias-González, Elisa Armenteros, Emilio José Laserna-Mendieta, Patricia Muñoz-Hernández, María Sendino, Javier Muñoz, Eren Efrem, Alfredo J Lucendo, Pedro Majano, Cecilio Santander

## Abstract

**Background:** The esophageal String Test (EST) is emerging as a minimally invasive alternative for assessing disease activity in eosinophilic esophagitis (EoE), yet the molecular composition of its protein content remains unknown.

**Objective:** To comprehensively characterize the esophageal EST proteome, define an EoE-associated luminal protein signature, and evaluate its biological and clinical relevance.

**Methods:** EST samples were obtained from patients with active EoE, healthy controls (C) and patients with gastroesophageal reflux disease (GERD). Label-free proteomic profiling was performed by liquid chromatography-tandem mass spectrometry. Differential protein accumulation, principal component analysis (PCA), pathway annotation, comparison with published esophageal tissue and salivary proteomes, receiver operating characteristic (ROC) analyses and correlations with clinical disease activity were performed.

**Results:** More than 3,000 proteins were identified in esophageal EST samples. Global proteomic profiling clearly separated patients with EoE from both C and GERD, whereas only limited differences were observed between the latter two groups. This enabled the identification of an EoE-associated luminal protein signature comprising inflammatory, epithelial, and tissue remodeling proteins. Comparison with published proteomic datasets indicated that the EST samples a complex esophageal luminal protein microenvironment containing proteins represented in esophageal mucosa and saliva, together with inflammatory and extracellular components. Several individual proteins showed high discriminatory performance in this exploratory cohort for distinguishing EoE from non-EoE subjects, and multiple proteins correlated with eosinophil counts and endoscopic measures of disease activity.

**Conclusions:** This study demonstrates that the EST provides access to a complex esophageal luminal protein microenvironment and establishes a molecular framework for its minimally invasive characterization in EoE.

## 1. INTRODUCTION

Eosinophilic esophagitis (EoE) is a chronic immune-mediated inflammatory disease characterized by symptoms of esophageal dysfunction and eosinophil-predominant inflammation of the esophageal mucosa ^1^. Current diagnostic criteria are based on symptoms of esophageal dysfunction together with a minimum of 15 eosinophils per high-power field (hpf) in esophageal biopsies after excluding alternative causes of esophageal eosinophilia. ^2,3^. Over the past two decades, both the incidence and prevalence of EoE have increased substantially worldwide ^4^, making this condition one of the leading causes of dysphagia and food impaction in children and adults and imposing a substantial and growing economic burden on healthcare systems ^5^ . Beyond eosinophilic inflammation, EoE is characterized by epithelial barrier dysfunction and tissue remodeling and may progress to fibrostenotic complications that ultimately lead to esophageal narrowing and strictures ^6–8^.

Assessment of disease activity currently relies on upper gastrointestinal endoscopy with esophageal biopsies ^2,3^. Although histological evaluation remains the diagnostic and monitoring gold standard, repeated endoscopic procedures are invasive, costly and burdensome for patients, often requiring sedation or anesthesia, particularly in the pediatric population. These limitations are especially relevant given the chronic relapsing nature of EoE, which necessitates repeated assessments to monitor treatment response and long-term disease control. Consequently, considerable efforts are currently directed towards developing minimally invasive approaches capable of accurately reflecting esophageal disease activity ^9,10^. The limited specificity of blood-based biomarkers currently investigated for EoE hampers their ability to distinguish EoE from coexisting atopic conditions, underscoring the potential of minimally invasive methods that capture disease-associated proteins directly at the site of disease^11^.

Among these approaches, the Esophageal String Test (EST) has emerged as one of the most promising technologies ^11^. The EST consists of a swallowed string that samples material present at the esophageal luminal surface during its residence in the esophagus. Initially explored as a minimally invasive approach to characterize the esophageal microbiome ^12^, its subsequent application in EoE has focused predominantly on protein biomarkers ^13,14^. Pioneering studies by Furuta and colleagues demonstrated that eosinophil-derived proteins recovered from the EST strongly correlate with esophageal eosinophil counts, eosinophil granule protein deposition, and disease activity. These studies provided early evidence that material recovered from the esophageal lumen reflects ongoing mucosal inflammation ^13^. These observations were subsequently confirmed in a multicenter study showing that the EST accurately discriminates active from inactive EoE in both children and adults, while correlating with histological and endoscopic activity ^14^. More recently, the application of the EST has been extended to treatment monitoring ^15^ and the evaluation of remodeling-associated biomarkers, including periostin, further supporting its clinical utility as a minimally invasive tool for disease assessment ^16^. Collectively, these studies support the EST as a valuable platform for assessing disease activity in EoE. However, they focused primarily on predefined candidate biomarkers, chiefly eosinophil-derived proteins and selected remodeling markers. As a result, the global molecular composition of the material sampled by the EST remains largely unknown. Whether the EST captures broader biological processes involved in EoE pathogenesis, including epithelial barrier dysfunction, epithelial differentiation, tissue remodeling and inflammatory pathways beyond eosinophil activation, has not been systematically investigated. Likewise, comparative analyses of the EST proteome in clinically relevant differential diagnoses, such as gastroesophageal reflux disease (GERD), remain scarce. Defining the molecular landscape within EST samples is therefore a critical step towards understanding the biological information captured by this technology and identifying new biomarker candidates.

Recent advances in mass spectrometry-based proteomics have substantially expanded our understanding of EoE biology ^17,18^. Using unbiased label-free proteomic profiling, we previously identified extensive alterations in the esophageal epithelial proteome involving eosinophilic inflammation, epithelial differentiation, keratinization, barrier integrity and extracellular matrix remodeling ^17,18^. Subsequently, we demonstrated that successful proton pump inhibitor (PPI) therapy largely reverses this molecular signature, while also identifying candidate proteins associated with treatment response ^19^. These findings highlight the value of global proteomic profiling for characterizing disease-associated molecular processes and identifying candidate biomarker.

In the present study, we applied unbiased label-free proteomic profiling to comprehensively characterize EST-derived luminal samples obtained from patients with active EoE, GERD and healthy controls (C). Specifically, we sought to define the molecular composition of the material sampled by the EST, identify the biological processes represented within the luminal proteome, evaluate the relationship between EST-derived proteins and clinical, endoscopic and histological measures of disease activity, and compare the EST proteome with previously characterized esophageal tissue and salivary proteomes. Ultimately, our goal was to determine whether the EST captures molecular information beyond established candidate biomarkers and to establish its potential as a minimally invasive platform for the molecular characterization of EoE.

## 2. MATERIALS AND METHODS

### Subjects

Adult patients with EoE were prospectively recruited from two Spanish hospitals— Hospital Universitario de La Princesa (Madrid) and Hospital General de Tomelloso (Ciudad Real)—between January 2024 and March 2025. The diagnosis of EoE was established according to evidence-based guidelines. Three esophageal biopsies were obtained from both the distal and proximal esophagus. Esophageal eosinophilia was defined as ≥15 eosinophils per hpf (corresponding to an area of 0.24 mm²) in one or more biopsy specimens. Patients with EoE were either treatment-naïve or had received previous therapy; however, all samples included in the present study were obtained during histologically active disease (≥15 eosinophils/hpf). Post-treatment samples were not included in the present analysis. Samples collected after initiation of treatment were not included in the present analysis and were reserved for subsequent longitudinal studies. Clinical symptoms were assessed using the Dysphagia Symptoms Score (DSS). Patients with GERD presented with recurrent symptoms consistent with the disease, mainly heartburn and regurgitation, and were frequently accompanied by dysphonia. In half of these patients, the diagnosis was based on clinical presentation and upper gastrointestinal endoscopy, whereas the remainder were confirmed via esophageal functional studies, including high-resolution esophageal manometry and ambulatory reflux monitoring.

Healthy controls were recruited from volunteers with no history of esophageal or gastrointestinal disorders, and who lacked symptoms suggestive of EoE or GERD, including dysphagia, food impaction, heartburn, or regurgitation.

This study (PI24/00082) was approved by the Research Ethics Committee of Instituto de Investigación Sanitaria Hospital Universitario de La Princesa (registry number 5840 January 2025). All patients and controls provided written informed consent prior to participation.

### Esophageal String Test sampling and protein extraction

The esophageal String Test (EST) was performed after an overnight fast using the Coiled Oesophageal String Test (CEST) developed at University Hospital Southampton (Southampton, UK). The CEST consists of a specially designed, coiled rayon string that is swallowed directly with water, eliminating the need for a dissolvable capsule. Participants were provided with a 500-mL bottle of water and instructed to drink *ad libitum* to facilitate device ingestion. The proximal end of the string was secured to the cheek with adhesive tape. The string remained *in situ* for 30 min to allow contact with the esophageal luminal surface before being gently withdrawn through the mouth.

Following retrieval, the gastric portion of the string was identified based on pH measurement and discarded. An approximately 20–22 cm segment corresponding to the portion exposed to the esophageal lumen was then selected for analysis, with minor adjustments according to participant height. The oral portion of the string was likewise discarded. Proteomic analyses were therefore restricted to the esophageal segment, minimizing direct contamination from the oral cavity. The selected segment was placed in 200 μL of extraction buffer containing 2% SDS and 100 mM HEPES and incubated at 95°C for 15 min to facilitate protein elution from the string. The resulting protein extracts were aliquoted and stored at −80°C until proteomic analysis.

### Mass spectrometry and statistical analysis

Extracted proteins were reduced with 15 mM TCEP, and alkylated with 30 mM CAA. Protein clean-up and digestion were executed via the SP3 methodology, inducing protein aggregation with 70% ethanol; following successive washes, overnight digestion was performed at 37 °C utilizing a LysC and trypsin mixture (1:50 w/w) in 50 mM ammonium bicarbonate. The reaction was quenched with TFA (1% final concentration), and the resulting peptides were desalted using C18 StageTips prior to reconstitution in 0.1% formic acid. LC-MS/MS analysis was conducted on a Vanquish Neo UHPLC system coupled to an Orbitrap Exploris 480 mass spectrometer utilizing a trap-and-elute configuration (Acclaim PepMap 100 C18 trap column and EASY-Spray PepMap RSLC C18 analytical column maintained at 50 °C). Chromatographic separation was achieved over a 60-minute effective gradient at a 250 nL/min flow rate. Data-independent acquisition (DIA) was employed, acquiring MS1 spectra across a 400–1000 m/z range, while MS/MS fragmentation was performed via HCD (NCE 30) following precursor ion isolation with 10 m/z overlapping windows at a spectral resolution of 15,000. DIA data processing was executed using DIA-NN (v1.8.1) via a library-free approach relying on deep learning predictions, searching against the Homo sapiens UniProt reference proteome. Enzyme specificity was restricted to Trypsin/P (allowing up to one missed cleavage), specifying cysteine carbamidomethylation as a fixed modification, alongside methionine oxidation and protein N-terminal acetylation as variable modifications; the false discovery rate (FDR) was strictly controlled at 1% with the Match Between Runs (MBR) algorithm enabled. Quantitative and statistical analyses were performed in Prostar, where, following valid value filtering, intensities were log2-transformed and normalized utilizing the LOESS method. Missing value imputation was stratified by missingness type: the SLSA algorithm for partially observed values (POV) and DetQuantile for values missing on an entire condition (MEC). Finally, differential protein abundance was evaluated employing empirical Bayes statistics (limma R package), establishing statistical significance based on predefined absolute log2 fold-change thresholds and an adjusted P-value < 0.05 (Benjamini-Hochberg correction).

## 3. RESULTS

### 3.1. Characteristics of EoE, GERD and healthy controls patients

The clinical, demographic, endoscopic and histological characteristics of the study population are summarized in Table 1. A total of 34 individuals were included: 17 patients with EoE, 8 patients with GERD and 9 healthy C. The groups showed comparable sex distribution and anthropometric characteristics, whereas patients with GERD were significantly older than both C and patients with EoE. As expected, dysphagia and food impaction were predominantly observed in patients with EoE, whereas heartburn and regurgitation were more frequent in the GERD group. Atopic disease was also more frequent in patients with EoE than in the other groups, although the difference between EoE and GERD did not reach statistical significance.

**Table 1.** Demographic, clinical, endoscopic and histological characteristics of the study population. Continuous variables are presented as mean ± SD and categorical variables as n (%). Endoscopic findings were assessed using the EoE Endoscopic Reference Score (EREFS), including inflammatory and fibrostenotic features. Histological disease activity in patients with EoE was assessed by total eosinophil counts and the EoE Histology Scoring System (EoEHSS), including grade (severity) and stage (extent) scores. Pairwise comparisons were performed using Mann–Whitney U tests for continuous variables and Fisher’s exact tests for categorical variables. P-values for pairwise comparisons are reported in the three rightmost columns. Abbreviations: C, control; DSS, Dysphagia Symptoms Score; EoE, eosinophilic esophagitis; EoEHSS, Eosinophilic Esophagitis Histology Scoring System; EREFS, EoE Endoscopic Reference Score; GERD, gastroesophageal reflux disease; HPF, high-power field; SD, standard deviation.

| Variable | Control (n = 9) | GERD (n = 8) | EoE (n = 17) | Control vs GERD | Control vs EoE | GERD vs EoE |
| --- | --- | --- | --- | --- | --- | --- |
| **Demographic characteristics** |  |  |  |  |  |  |
| Age (years), mean ± SD | 37.0 ± 11.5 | 52.5 ± 13.7 | 31.5 ± 9.9 | 0.034 | 0.305 | <0.001 |
| Male sex, n (%) | 7 (77.8) | 5 (62.5) | 13 (76.5) | 0.620 | 1.000 | 0.640 |
| **Symptoms** |  |  |  |  |  |  |
| Dysphagia, n (%) | 0 (0.0) | 3 (37.5) | 17 (100.0) | 0.082 | <0.001 | 0.001 |
| Food impaction, n (%) | 0 (0.0) | 0 (0.0) | 12 (70.6) | 1.000 | <0.001 | 0.002 |
| Heartburn, n (%) | 1 (11.1) | 8 (100.0) | 9 (52.9) | <0.001 | 0.087 | 0.026 |
| Regurgitation, n (%) | 0 (0.0) | 8 (100.0) | 8 (47.1) | <0.001 | 0.023 | 0.022 |
| DSS score, mean ± SD | — | — | 9.9 ± 3.8 | — | — | — |
| **Atopic diseases** |  |  |  |  |  |  |
| Atopic disease, n (%) | 3 (33.3) | 3 (37.5) | 14 (82.4) | 1.000 | 0.028 | 0.061 |
| Food allergy, n (%) | 0 (0.0) | 1 (12.5) | 6 (35.3) | 0.471 | 0.063 | 0.362 |
| **Endoscopic findings** |  |  |  |  |  |  |
| Total EREFS score, mean ± SD | — | 0.1 ± 0.4 | 3.7 ± 1.5 | — | — | <0.001 |
| Fibrostenotic EREFS score, mean ± SD | — | 0.1 ± 0.4 | 1.2 ± 1.0 | — | — | 0.006 |
| Inflammatory EREFS score, mean ± SD | — | 0.0 ± 0.0 | 2.5 ± 1.1 | — | — | <0.001 |
| **Histological findings** |  |  |  |  |  |  |
| Total eosinophils, mean ± SD | — | — | 99.4 ± 85.8 | — | — | — |
| Grade Score, mean ± SD | — | — | 0.7 ± 0.3 | — | — | — |
| Stage Score, mean ± SD | — | — | 0.7 ± 0.3 | — | — | — |

Abbreviations: EoE, eosinophilic esophagitis; GERD, gastroesophageal reflux disease; EREFS, EoE Endoscopic Reference Score; HPF, high-power field; DSS, diagnostic score/system as defined in the study.
Notes: Continuous variables are shown as mean ± SD. Categorical variables are shown as n (%). Pairwise comparisons were performed using Mann-Whitney U tests for continuous variables and Fisher's exact tests for categorical variables.

Endoscopic assessment revealed substantially greater disease severity in patients witg EoE than in those with GERD, as evidenced by higher total EREFS scores ^20^ and significant differences involving both inflammatory and fibrostenotic features (Table 1). Histological assessment confirmed active eosinophilic inflammation in the EoE group, with a mean peak eosinophil count of 63.9 ± 45.9 eosinophils/hpf. EoE severity and extent were further characterized histologically using the EoE Histology Scoring System (EoEHSS) ^21^, with mean final grade and stage scores of 0.7 ± 0.3.

### 3.2. Global String Test proteomic profiling reveals a distinct EoE-associated profile

Mass spectrometry analysis of EST samples identified a total of 4,030 proteins, of which 3,049 passed quality filtering and were retained for subsequent analyses. This extensive proteome coverage illustrates the molecular complexity of the material recovered by the EST and provides the basis for an unbiased comparison of the proteomic profiles of C, patients with GERD and patients with EoE.

To determine whether the global proteomic profile differed among diagnostic groups, unsupervised principal component analysis (PCA) was performed using all quantified proteins. Samples from patients with EoE showed a clear shift from both C and patients with GERD, whereas the latter two groups exhibited substantial overlap (Figure 1A). Thus, group separation emerged at the whole-proteome level without prior selection of disease-associated proteins, indicating broad differences in the protein composition recovered by the EST in EoE compared with the other two groups.

**Figure 1.**
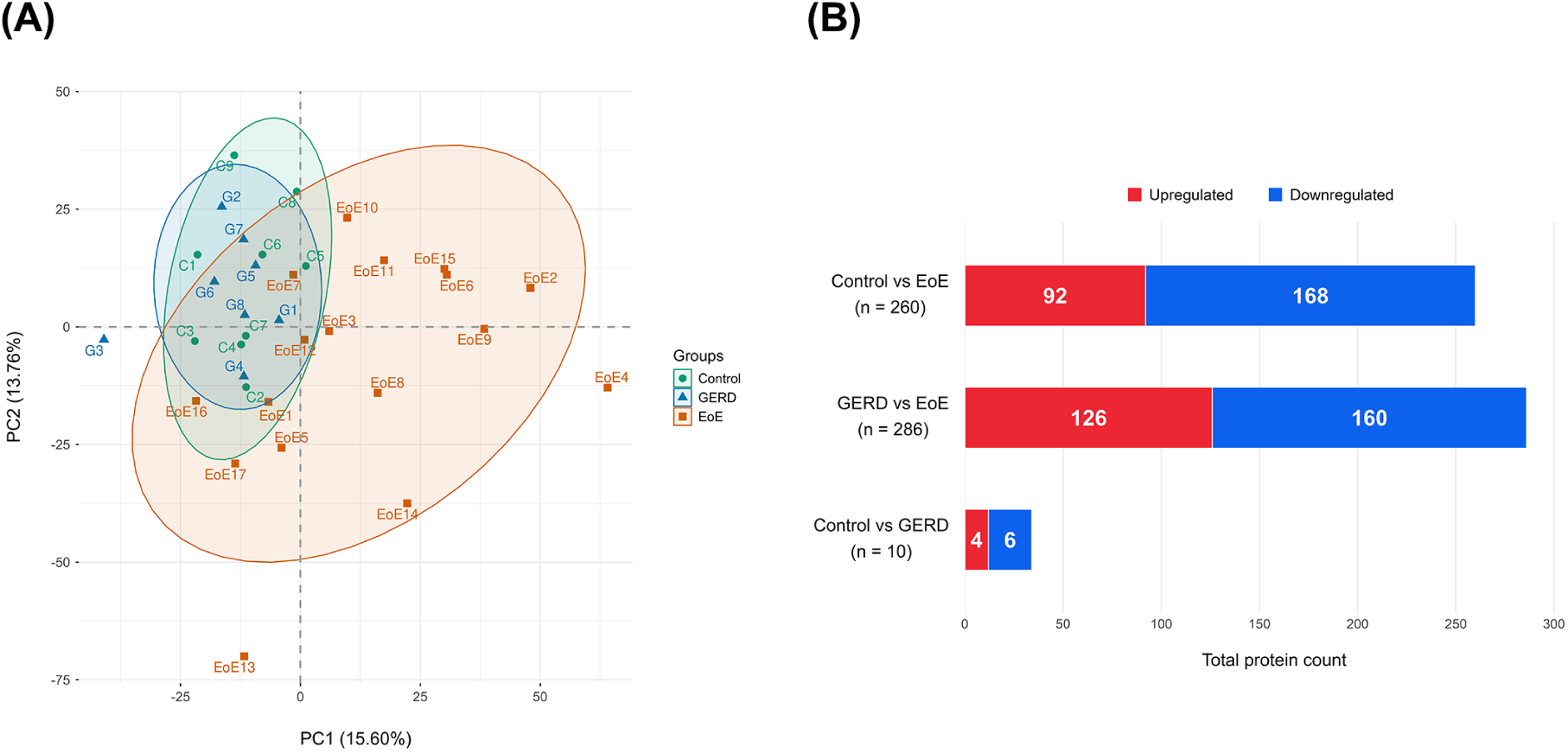
Global proteomic profiling of esophageal String Test samples distinguishes eosinophilic esophagitis from healthy controls and GERD. (A) Unsupervised principal component analysis (PCA) based on all quantified proteins showing the global distribution of healthy controls (C), patients with gastroesophageal reflux disease (GERD), and patients with eosinophilic esophagitis (EoE). (B) Number of differentially accumulated proteins (DAPs) identified in each pairwise comparison using an adjusted P ≤ 0.05. Red and blue indicate proteins with increased and decreased abundance, respectively, in the target condition relative to the first-named reference group (e.g., EoE relative to C in the C vs EoE comparison). Abbreviations: C, controls; DAPs, differentially accumulated proteins; EoE, eosinophilic esophagitis; GERD, gastroesophageal reflux disease; PCA, principal component analysis.

Differential protein abundance analysis further supported these observations. Using an adjusted P-value (FDR) < 0.05 as the sole criterion for differential accumulation, without applying a fold-change threshold at this stage, 260 proteins were identified as DAPs between C and EoE, including 92 proteins increased and 168 decreased in EoE. Similarly, 286 DAPs were identified between GERD and EoE, with 126 proteins increased and 160 decreased in EoE (Figure 1B). Remarkably, only 10 proteins differed between C and GERD (4 increased and 6 decreased), indicating that the EST proteomic profiles of the two non-EoE groups were largely comparable. PCA, hierarchical clustering/heatmaps and volcano plots for the individual pairwise comparisons are shown in Supplementary Figures S1–S3, respectively.

Collectively, both the global proteomic distribution and differential abundance analyses demonstrated that EoE was associated with extensive alterations of the EST proteome compared with either C or GERD, whereas differences between the two non-EoE groups were minimal.

### 3.3. Definition of an EoE-associated esophageal luminal protein signature

As shown in Figure 1, the similarity between C and patients with GERD provided the rationale for combining both groups into a single non-EoE reference group to define an EoE-associated protein signature (Figure 2). Differential protein abundance was assessed between EoE and non-EoE subjects, and an additional fold-change threshold (|log₂FC| ≥ 0.585, corresponding to a ≥1.5-fold difference), together with an adjusted P ≤ 0.05, was applied to prioritize proteins showing both statistically significant and substantial differences in abundance. This selection yielded 152 proteins, including 40 with increased and 112 with decreased abundance in EoE, collectively defining the EoE-associated EST protein signature (Figure 2A).

**Figure 2.**
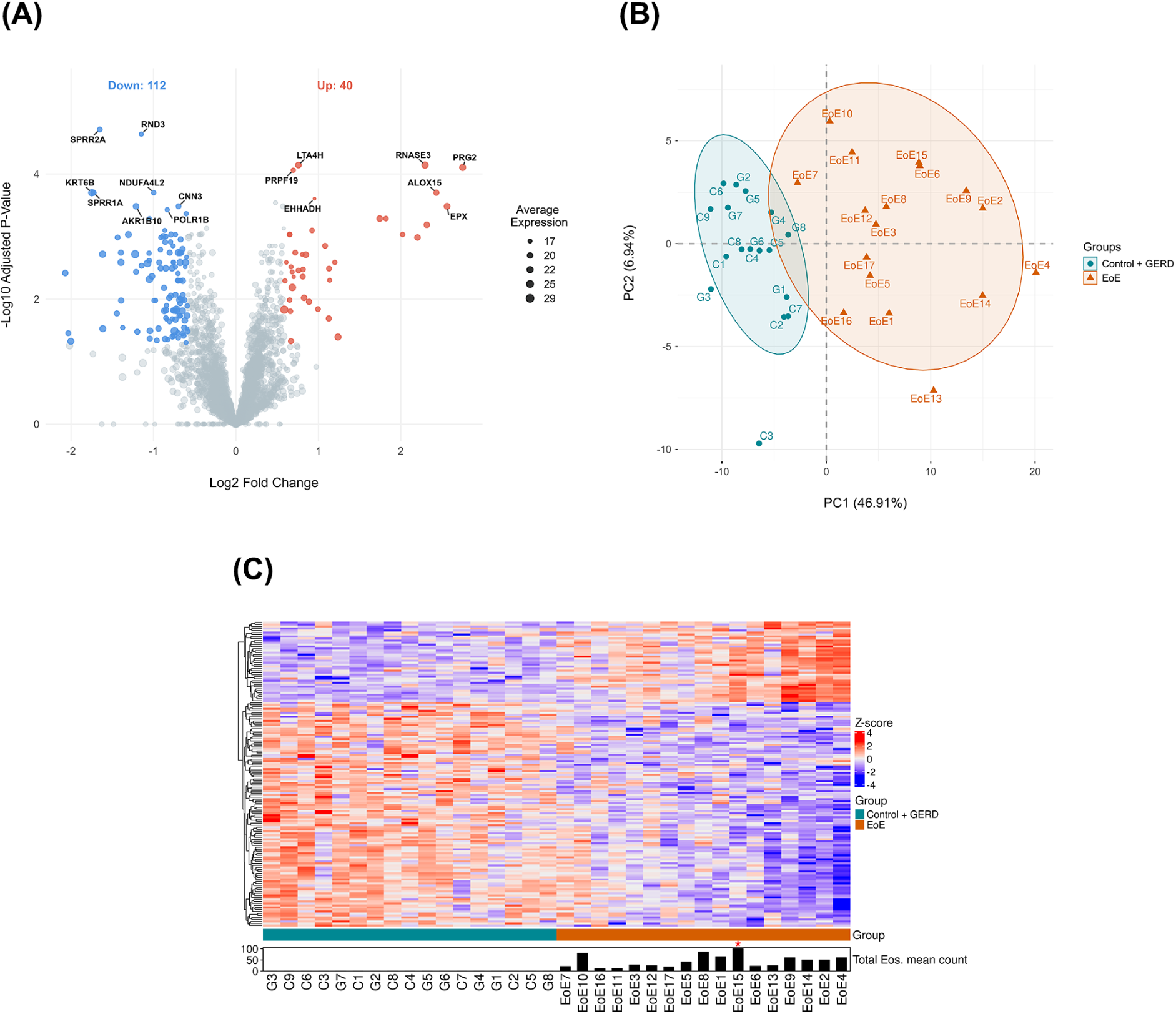
Definition of an EoE-associated protein signature in the esophageal String Test. (A) Volcano plot showing differentially accumulated proteins (DAPs; n = 152) between patients with EoE and the combined non-EoE group (C + GERD). Thresholds of adjusted P ≤ 0.05 and |log2FC| ≥ 0.585 were applied. Red and blue dots indicate proteins with increased (n = 40) and decreased (n = 112) abundance in EoE, respectively. The 15 proteins with the lowest adjusted P-values are highlighted. Dot size corresponds to average protein expression levels. (B) Principal component analysis (PCA) performed using the proteins included in the EoE-associated luminal protein signature, showing separation between EoE and the combined non-EoE group. (C) Heatmap representing the abundance, after z-score normalization, of proteins comprising the EoE-associated luminal protein signature across all study samples. Proteins were hierarchically clustered using Euclidean distance and Ward’s linkage. Samples were stratified by group and sorted along a molecular gradient based on their Euclidean distance to the mean protein expression profile of the opposing group. Bottom annotations indicate mean total eosinophil counts, with asterisks denoting values exceeding 100 eosinophils. Abbreviations: C, controls; DAPs, differentially accumulated proteins; EoE, eosinophilic esophagitis; GERD, gastroesophageal reflux disease; log2FC, log2 fold change; PCA, principal component analysis.

The distribution of this 152-protein signature across individual samples was subsequently examined using complementary multivariate approaches. PCA based exclusively on these proteins showed clear separation of EoE from the combined C+GERD group (Figure 2B). Consistently, hierarchical clustering largely segregated EoE from non-EoE samples and revealed coordinated patterns of increased and decreased protein abundance (Figure 2C). These findings illustrate the distinct molecular profile captured by the EoE-associated EST signature and provide the basis for subsequent functional characterization of the luminal proteome.

### 3.4. The String Test captures a complex esophageal luminal protein microenvironment

To characterize the biological composition of the esophageal luminal protein microenvironment sampled by the EST, we compared the complete EST proteome with reference proteomes from human esophageal mucosa, whole saliva, our previously published EoE biopsy dataset, and an independent EoE biopsy proteome reported by Rochman et al ^18,22–24^. Of the 3,049 EST proteins, 2,348 were also detected in the human esophageal mucosal proteome, 1,315 in whole saliva, 1,412 in our EoE biopsy dataset, and 1,701 in the independent EoE biopsy dataset (Figure 3A). These findings indicate that the EST samples a complex luminal protein microenvironment containing proteins represented in both esophageal tissue and saliva.

We next examined the distribution of the 152-protein EoE-associated signature across the same reference proteomes. Of these proteins, 109 (71.71%) were detected in the human esophageal mucosal proteome, compared with 70 (46.05%) in whole saliva, 67 (44.08%) in our EoE biopsy proteome, and 82 (53.95%) in the independent EoE biopsy dataset (Figure 3B). Thus, the EoE-associated signature showed its greatest representation in the esophageal mucosal reference proteome, supporting a close relationship between the disease-associated luminal signal detected by the EST and molecular alterations present in the esophageal mucosa.

### 3.5. Functional characterization of the EoE-associated protein signature

To gain biological insight into the EoE-associated protein signature, functional enrichment analysis was performed separately for proteins increased and decreased in EoE, revealing a marked functional asymmetry between both components of the signature (Figure 4). Proteins increased in EoE were predominantly enriched in immune-related processes, including lymphocyte-mediated immunity, leukocyte-mediated cytotoxicity, T-cell-associated responses and innate immune responses at mucosal surfaces. In contrast, proteins decreased in EoE showed coherent enrichment in epithelial programs related to epidermal and epithelial development, keratinocyte differentiation, keratinization, intermediate filament organization and epithelial cell–cell adhesion.

**Figure 4.**
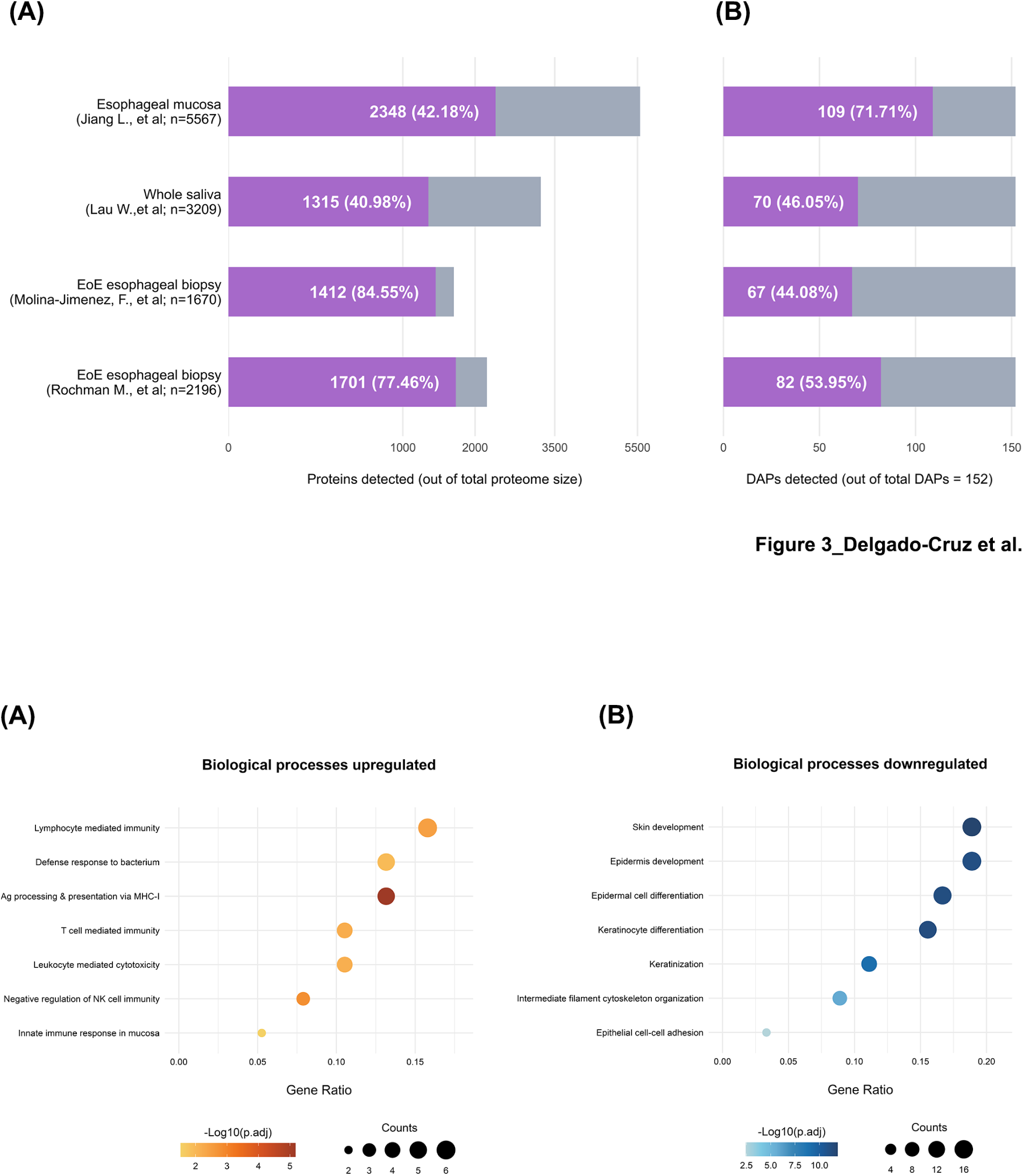
Functional characterization of the EoE-associated luminal protein signature. (A) Enriched Gene Ontology (GO) biological processes associated with proteins increased in EoE, predominantly involving immune-related processes. (B) Enriched GO biological processes associated with proteins decreased in EoE, mainly related to epithelial and epidermal development, keratinocyte differentiation and keratinization. Dot size represents the number of proteins associated with each biological process, and color intensity reflects statistical significance, expressed as −log10(adjusted P-value). Abbreviations: EoE, eosinophilic esophagitis; EST, esophageal String Test; GO, Gene Ontology.

The inflammatory component included several eosinophil-associated proteins, including eosinophil peroxidase (EPX; EPO), Charcot–Leyden crystal protein/galectin-10 (CLC; GAL10), proteoglycan 2/eosinophil major basic protein 1 (PRG2; MBP-1), proteoglycan 3/eosinophil major basic protein 2 (PRG3; MBP-2), ribonuclease A family member 2/eosinophil-derived neurotoxin (RNASE2; EDN), and ribonuclease A family member 3/eosinophil cationic protein (RNASE3; ECP), together with additional proteins involved in innate and adaptive immune responses. Conversely, proteins decreased in EoE included multiple components associated with squamous epithelial differentiation and cornification, including envoplakin (EVPL), small proline-rich proteins (SPRR1A, SPRR2A and SPRR3) and keratin 6B (KRT6B), as well as proteins involved in epithelial adhesion, barrier organization and cytoskeletal architecture, such as plakophilin 3 (PKP3), catenin beta-1 (CTNNB1).

Together, these findings show that the EoE-associated luminal protein signature combines increased immune and eosinophil-associated proteins with decreased proteins related to epithelial differentiation, keratinization and structural organization, indicating that the EST captures both inflammatory and epithelial alterations associated with active EoE.

### 3.6. Clinical associations and diagnostic performance of the String Test protein signature

To investigate the clinical relevance of the EoE-associated protein signature, we examined its relationship with established measures of disease activity. To limit the number of comparisons while maintaining a selection independent of clinical variables, the 15 proteins with the largest fold changes in each direction (15 increased and 15 decreased in EoE) were selected from the 152-protein signature. Associations with endoscopic and histological parameters were assessed using Spearman’s rank correlation analysis (Figure 5A).

**Figure 5.**
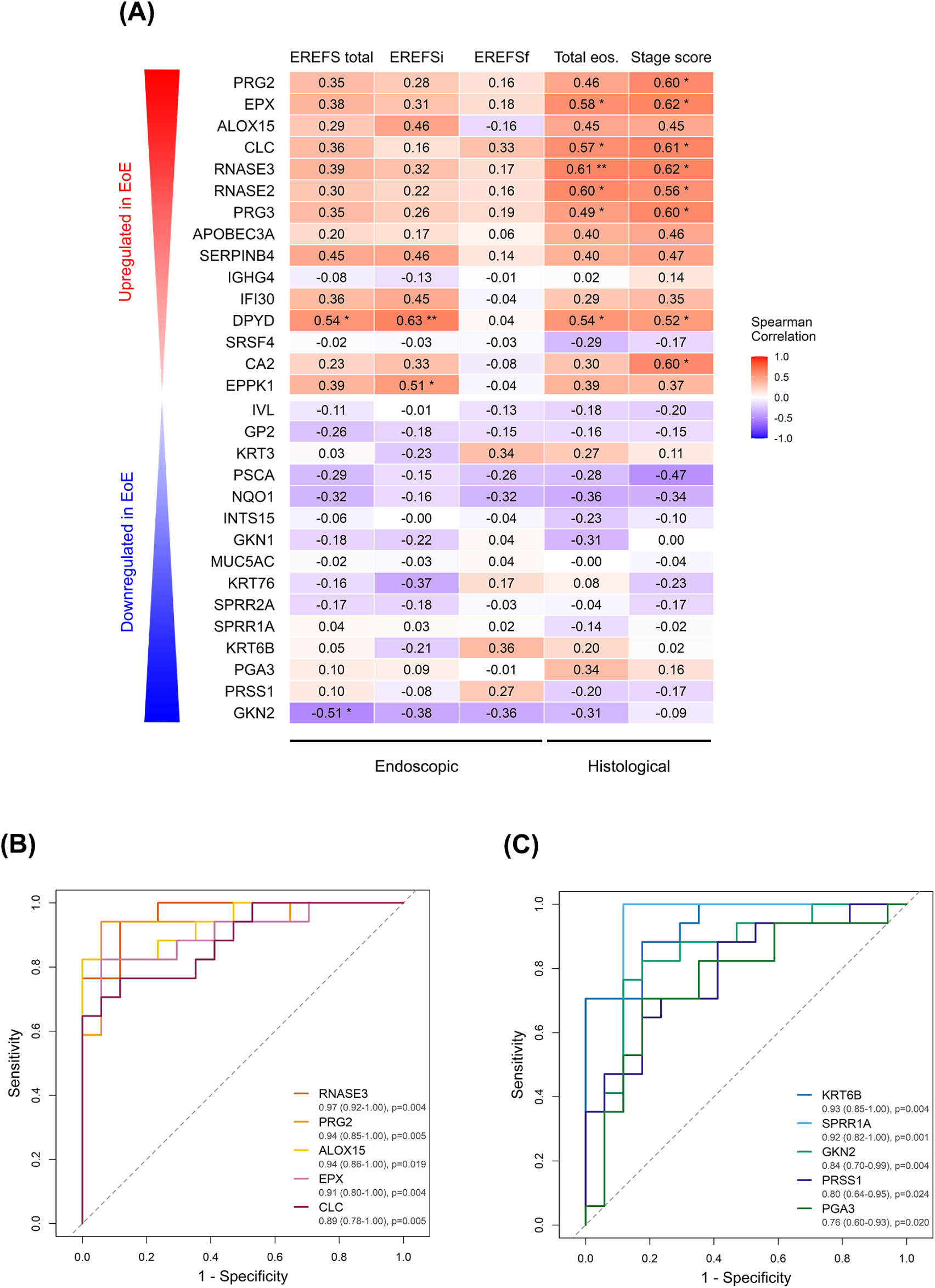
Association of the EoE-associated esophageal luminal protein signature with disease activity and discriminatory performance. (A) Spearman correlation matrix between the 15 most strongly increased and 15 most strongly decreased proteins of the EoE-associated signature, selected according to fold-change magnitude, and endoscopic and histological measures of disease activity. Endoscopic parameters included total EREFS and its inflammatory (EREFSi) and fibrostenotic (EREFSf) components; histological parameters included total eosinophil counts and EoEHSS stage score. Color intensity represents the Spearman correlation coefficient (ρ), with positive correlations shown in red and negative correlations in blue. Asterisks indicate statistically significant correlations. (B–C) Receiver operating characteristic (ROC) curves evaluating discrimination between EoE and non-EoE subjects (C and GERD) for the five proteins showing the largest fold changes among proteins increased (B) or decreased (C) in EoE. AUC values with 95% confidence intervals and p values are shown. Abbreviations: AUC, area under the curve; C, controls; EoE, eosinophilic esophagitis; EoEHSS, Eosinophilic Esophagitis Histology Scoring System; EREFS, EoE Endoscopic Reference Score; EREFSf, fibrostenotic EREFS; EREFSi, inflammatory EREFS; GERD, gastroesophageal reflux disease; ROC, receiver operating characteristic.

Proteins increased in EoE showed predominantly positive associations with measures of disease activity. Eosinophil-associated proteins, including PRG2, EPX, CLC, RNASE3, RNASE2 and PRG3, were particularly associated with histological activity, with several showing moderate-to-strong correlations with eosinophil counts and EoEHSS stage score. Associations with endoscopic severity were also observed, although these were more heterogeneous. Conversely, proteins decreased in EoE tended to show inverse or weaker associations with disease activity. Overall, these findings indicate that components of the EoE-associated luminal protein signature reflect complementary histological and endoscopic features of active disease.

We next evaluated the discriminatory performance of proteins showing the largest changes in EoE. ROC analysis of the five most strongly increased proteins showed high discrimination between EoE and non-EoE subjects, with RNASE3, PRG2, ALOX15, EPX and CLC yielding AUC values ranging from 0.90 to 0.97 (Figure 5B). Among the five most strongly decreased proteins, KRT6B and SPRR1A showed the highest discriminatory performance (AUCs of 0.93 and 0.92, respectively), whereas GKN2, PRSS1 and PGA3 yielded AUC values of approximately 0.80–0.86 (Figure 5C). Together, these analyses link the EoE-associated EST protein signature to established measures of disease activity and identify individual proteins with strong discriminatory capacity for EoE.

## DISCUSSION

The present study provides the first comprehensive molecular characterization of the biological compartment sampled by the EST in EoE. Using label-free proteomic profiling, we identified more than 3,000 proteins, demonstrating that this minimally invasive device captures a remarkably complex protein landscape encompassing epithelial differentiation and barrier integrity, inflammation, extracellular matrix organization and tissue remodeling. Previous EST studies established its clinical utility by quantifying a limited number of candidate biomarkers, primarily eosinophil-derived granule proteins ^12,14–17^. In contrast, our results define the global molecular composition of the material recovered by the EST and provide a biological framework for interpreting EST-derived biomarkers. Importantly, major eosinophil-associated proteins previously investigated using the EST ^12,14–17^, including EPX, RNASE2, RNASE3, PRG2, PRG3 and CLC, were all significantly increased in EoE in our proteomic analysis. This provides internal validation of the analytical approach while placing these established biomarkers within a broader molecular pattern involving eosinophilic inflammation, epithelial differentiation and tissue remodeling.

A major finding of this study is the extensive overlap between the EST proteome and both esophageal tissue and salivary proteomes ^18,22–24^. Comparison with our previously published and independent EoE biopsy datasets demonstrated substantial overlap both across the global proteome and within the EoE-associated protein signature. Despite sampling the esophageal lumen rather than tissue, these findings indicate that the EST captures a substantial component of the molecular landscape represented in the esophageal mucosa. Although salivary proteins are also present and the precise origin of individual proteins cannot always be established, the extensive representation of proteins also detected in esophageal tissue argues against the recovered material merely reflecting saliva. Importantly, the EST may provide information beyond that obtained from the epithelial proteome alone. It recovers extracellular proteins, eosinophil granule products, secreted mediators and molecules associated with epithelial injury and tissue remodeling, thereby providing access to a luminal protein microenvironment reflecting epithelial, immune and extracellular components. Although the EST cannot provide information on tissue architecture or cellular localization, it may capture molecular information complementary to that obtained from conventional biopsies. Future studies dissecting the relative contribution of tissue, inflammatory and salivary compartments will help define the origin and biomarker potential of individual proteins.

The biological composition of the EoE-associated signature further supports this interpretation. Proteins involved in epithelial differentiation, keratinization, cell adhesion and barrier integrity were altered alongside eosinophil granule proteins and molecules associated with extracellular matrix organization and tissue repair ^18,25^. This coordinated pattern is consistent with the interconnected processes of epithelial dysfunction, type 2 inflammation and remodeling that characterize EoE pathogenesis. Thus, the EST captures multiple components of active simply reflecting eosinophilic inflammation.

Interestingly, IGHG4 was also increased in the EoE-associated EST signature and is consistent with the prominent local IgG4 response previously described in EoE^26^. Increased IgG4 levels, IgG4-positive plasma cells and extracellular IgG4 deposition have been reported in esophageal tissue, together with food-specific IgG4 responses. Tissue IgG4 has also been associated with eosinophilic inflammation and histological disease activity, whereas a comparable increase in IgE has not been observed^27^. Although the pathogenic role of IgG4 remains uncertain, its detection in the EST proteome provides further evidence that this luminal compartment reflects local immune responses associated with EoE.

The inclusion of patients with GERD provided a clinically relevant comparator for evaluating the specificity of the EST proteomic profile. Although GERD and EoE may share symptoms and some endoscopic and histological features ^28^, previous molecular studies have demonstrated distinct tissue transcriptomic and proteomic signatures in these conditions ^29,30^. In our study, only limited proteomic differences were observed between patients with GERD and C, in marked contrast to the extensive changes detected in EoE. This finding should be interpreted cautiously, as GERD-associated mucosal injury and inflammation may vary with disease activity and treatment status. Moreover, GERD-associated molecular alterations may be less prominently represented in the esophageal luminal proteome than the pronounced inflammatory and epithelial changes associated with active EoE.

Thus, the similarity between C and GERD EST proteomes should not be interpreted as evidence of biological equivalence between these conditions. Nevertheless, EoE exhibited a clearly distinct luminal proteomic profile compared with both groups, supporting the ability of EST proteomics to capture disease-associated molecular alterations in active EoE.

The association between EST proteins and established measures of disease activity further supports their clinical relevance. Eosinophil-associated proteins showed particularly strong relationships with histological activity, while the broader signature captured molecular components extending beyond eosinophilic infiltration alone ^14–17^. This multidimensional information may be particularly valuable for disease monitoring, since conventional biopsies provide a localized tissue snapshot and require repeated endoscopic procedures. By contrast, the EST samples proteins released into the esophageal lumen during device placement and may therefore integrate biological activity across a broader mucosal surface. Rather than replacing histology, EST proteomics could complement it by providing additional information on the molecular state of the esophageal mucosa.

Importantly, histological remission may not necessarily imply complete molecular resolution of EoE ^31^. Persistent transcriptomic and ^31,32^ and proteomic ^19^ abnormalities, including remodeling- and barrier-associated molecules, have been reported even in patients achieving deep histological remission. In this context, persistent detection of extracellular eosinophil granule proteins or remodeling-associated molecules in the esophageal lumen could reflect ongoing processes not fully captured by routine eosinophil enumeration ^12^. Conversely, normalization of luminal biomarkers might precede complete tissue recovery in some patients. Although these possibilities require prospective longitudinal validation, they highlight the potential of luminal proteomics to provide complementary information on disease resolution and tissue repair beyond conventional histopathological assessment.

A major strength of this study is its unbiased discovery-based approach, which enabled simultaneous characterization of more than 3,000 proteins rather than restricting the analysis to predefined candidate biomarkers. This provides a rational basis for developing future targeted panels representing complementary biological pathways. However, the study has several limitations. The sample size is relatively modest, and the results should therefore be independently validated in larger multicenter cohorts. Moreover, the cross-sectional design precludes evaluation of molecular changes during treatment, making longitudinal studies with serial EST sampling an important next step. Finally, label-free mass spectrometry provides robust relative quantification across biological samples but does not provide direct absolute quantification across different proteins.

Translation into clinical practice will therefore require validation of the most informative candidates using simplified targeted assays, such as immunoassays or targeted mass spectrometry, capable of establishing clinically relevant cut-off values.

The concept of the esophageal luminal protein microenvironment may also extend beyond the EST and EoE. Esophageal surface-sampling devices such as the Cytosponge have demonstrated the clinical value of minimally invasive sampling in Barrett’s esophagus^34^ and are being explored in EoE for assessing disease activity and facilitating patient monitoring^35^. Together, these approaches support the broader concept that esophageal surface sampling can provide clinically relevant molecular information without the need for conventional biopsy. Comprehensive proteomic profiling could further expand this information by integrating epithelial, extracellular and secreted components of the esophageal lumen, offering potential applications in other diseases characterized by extensive epithelial remodeling.

Pre-analytical factors should also be considered when interpreting luminal proteomic profiles. Detergent exposure has been shown to disrupt esophageal epithelial barrier function and promote eosinophilic inflammation^36^. More recently, a study using EST samples after toothbrushing with an SDS-containing toothpaste identified an acute response dominated by salivary and mucosal defense proteins, with minimal overlap with the EoE-associated signature identified here^37^. Together, these observations highlight the importance of controlling recent oral hygiene and other potential pre-analytical variables before EST sampling.

In conclusion, this study demonstrates that the EST captures a complex esophageal luminal protein microenvironment containing proteins represented in esophageal tissue together with extracellular and soluble components. Rather than serving simply as a minimally invasive alternative to tissue sampling, the EST enables characterization of a complementary biological compartment reflecting epithelial dysfunction, inflammation and tissue remodeling. This framework provides a strong rationale for developing minimally invasive molecular strategies for disease monitoring, biomarker discovery and therapeutic assessment in EoE and, potentially, other esophageal disorders.

## Financial support

PM and CS are supported by grants PI17/0008 and PI24/00082, from Instituto de Salud Carlos III (ISCIII, Spain) and co-funded by Fondo Europeo de Desarrollo Regional (FEDER). CS and EJ L-M are supported by grant PI21/00579 of ISCIII, Spain and co-funded by FEDER. FD-C is supported by the Comunidad de Madrid (Consejería de Educación, Ciencia y Universidades) through the ’Ayudas para la contratación de personal investigador predoctoral en formación’ program (PIPF-2024-SAL-GL-34151). JM is supported by grant PID2021-123144OB-I00 funded by MCIN /AEI /10.13039/501100011033 / FEDER, UE. EA is a recipient of an INVESTIGO contract from Ministry of Labour and Social Economy, the national public employment service (SEPE) (INVESTIGO Exp. 2022-C23.I01.P03. S0020-0000031.2) partly funded by the European Social Fund, NextGenerationEU, and Recovery, Transformation and Resilience Plan.

## Data availability statement

Additional datasets will be available upon request at time of publication. For inquiries, please contact the corresponding author.

## Data Availability

All data produced in the present study are available upon reasonable request to the authors

## Acknowledgments

We express our gratitude to Drs. Manuel Gómez and Miguel Sampedro-Nuñez for critical review of the manuscript.

## Author disclosure

### Conflict of interest statement

The authors declare no conflicts of interest.

### Author contributions

Conceived and designed the study: PM, CS

Participated in the clinical management of patients: SC, ER, VM-D, JF-P, PM-H, AJL, EE, CS

Performed the experiments: FD-C, FM-J, LA-G, EA, EJL-M, MM Analyzed and discussed the data: FD-C, FM-J, PM, CS

Wrote the paper: FD-C, FM-J, PM, CS

All the authors read, provided comments, and approved the final version of the manuscript.

### Abbreviations

CLC: Charcot–Leyden crystal protein
DAP: differentially accumulated protein
EoE: Eosinophilic esophagitis
EoEHSS: Eosinophilic Esophagitis Histology Scoring System
EREFS: EoE Endoscopic Reference Score
EPX: Eosinophil peroxidase
EST: Esophageal string test
GERD: Gastroesophageal reflux disease
C: Healthy controls
hpf: High-power field
PCA: Principal component analysis
RNASE: ribonuclease A family member
ROC: Receiver operating characteristic

---

**Figure 3.** The String Test captures the esophageal luminal protein microenvironment.

(A) Overlap between the 3,049 proteins identified in EST samples and reference proteomes from human esophageal mucosa, whole saliva, our previously published EoE esophageal biopsy dataset, and an independent EoE biopsy proteome. Bars indicate the number of shared proteins and their percentage relative to each reference proteome. (B) Representation of the 152-protein EoE-associated EST signature across the same reference proteomes. Bars indicate the number of signature proteins detected in each reference dataset and their percentage relative to the complete 152-protein signature. Abbreviations: EoE, eosinophilic esophagitis; EST, esophageal String Test.

**Supplementary Figure S1.**
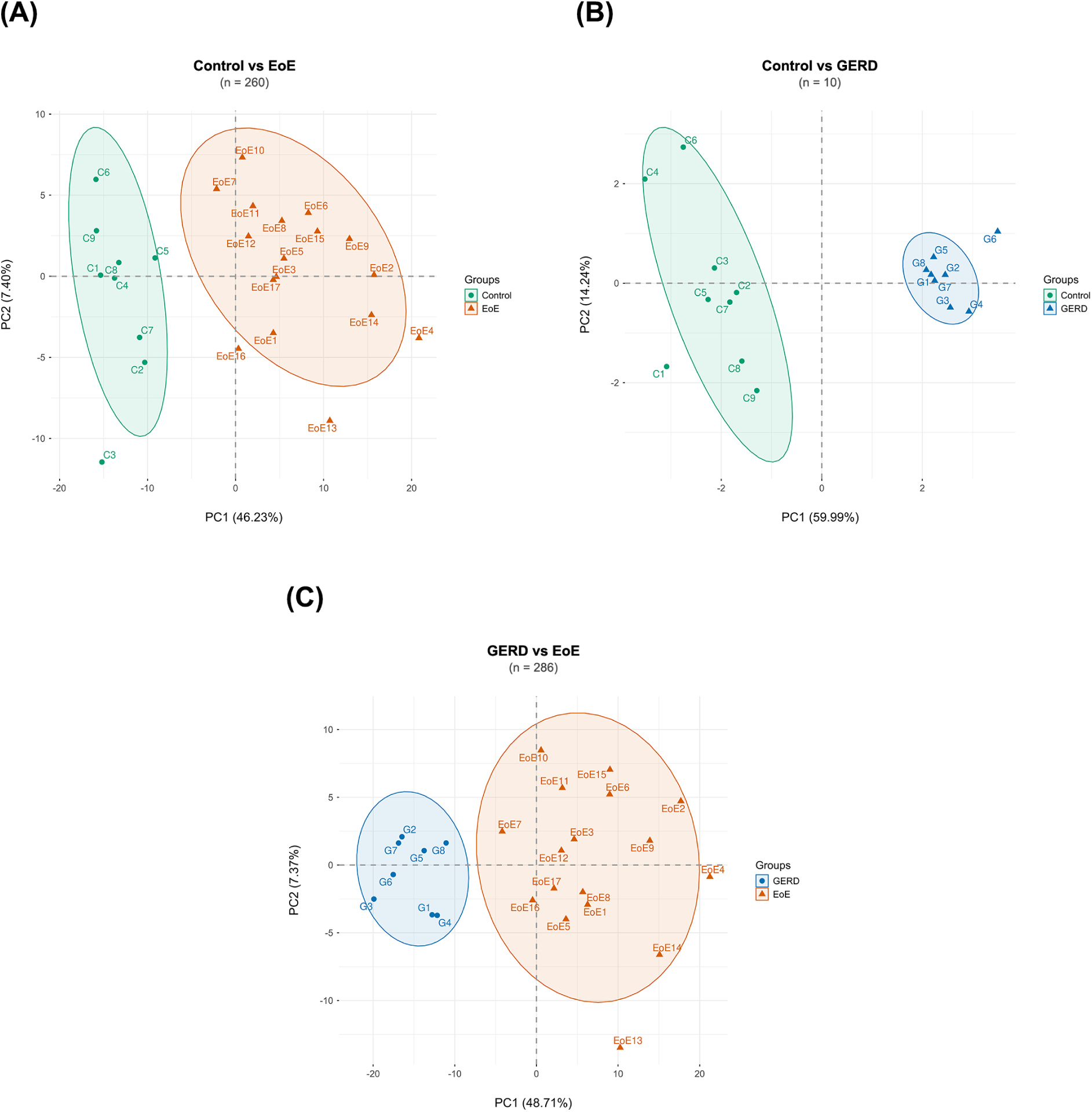
Principal component analysis (PCA) of individual pairwise comparisons. PCAfor each pairwise comparison, performed using the differentially accumulated proteins (DAPs) identified in the corresponding contrast: (A) C vs EoE, (B) C vs GERD, and (C) GERD vs EoE. Abbreviations: C, healthy controls; DAPs, differentially accumulated proteins; EoE, eosinophilic esophagitis; GERD, gastroesophageal reflux disease; PCA, principal component analysis

**Supplementary Figure S2.**
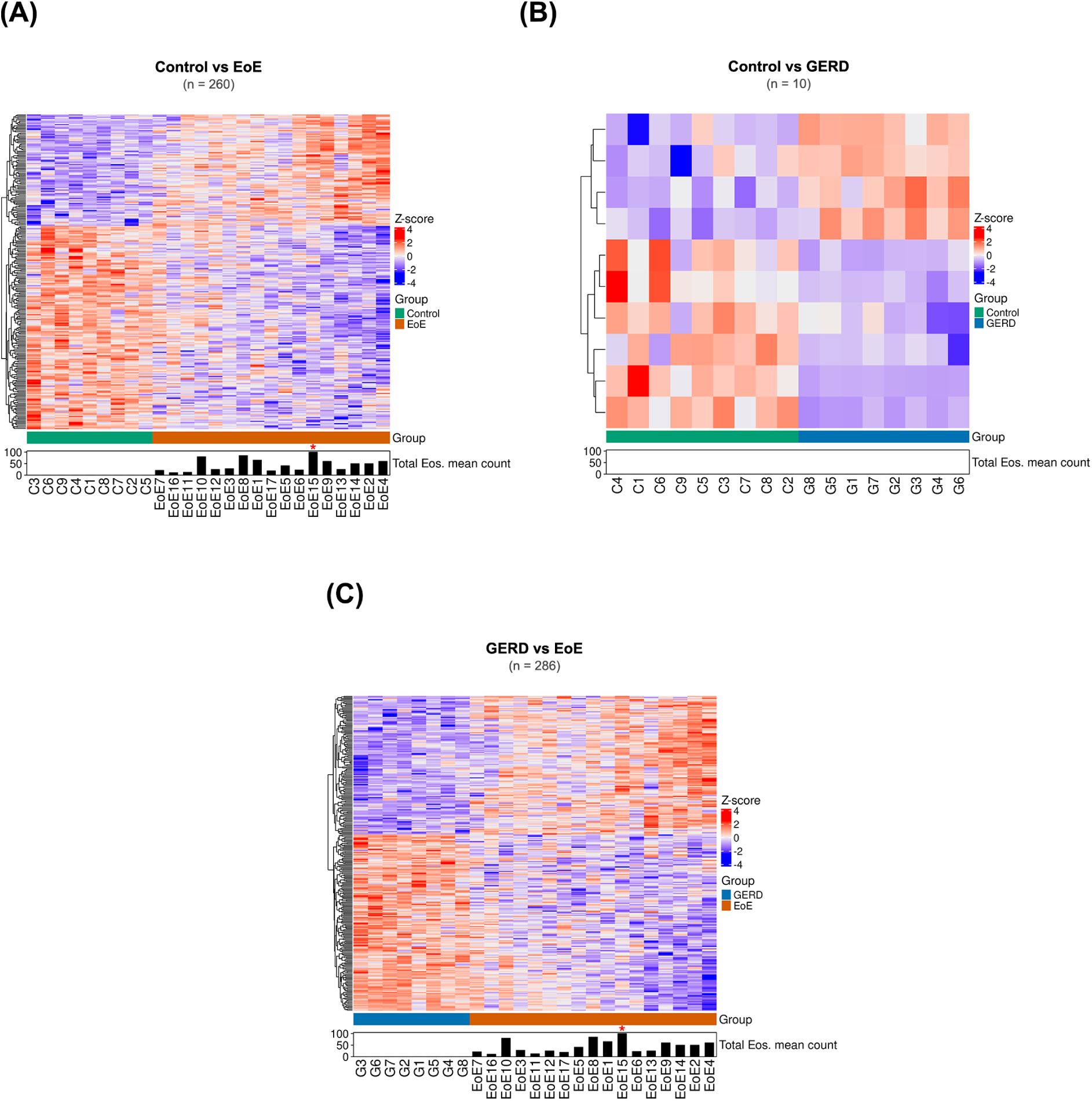
Heatmaps of differentially accumulated proteins in individual pairwise comparisons. Heatmaps representing protein abundance, after z-score normalization, of the differentially accumulated proteins (DAPs) identified in each pairwise comparison: (A) C vs EoE, (B) C vs GERD, and (C) EoE vs GERD. Proteins were hierarchically clustered using Euclidean distance and Ward’s linkage. Samples were stratified by group and sorted along a molecular gradient based on their Euclidean distance to the mean protein expression profile of the opposing group. Bottom annotations indicate mean total eosinophil counts, with asterisks denoting values exceeding 100 eosinophils. Abbreviations: C, Control; DAPs, differentially accumulated proteins; EoE, eosinophilic esophagitis; GERD, gastroesophageal reflux disease.

**Supplementary Figure S3.**
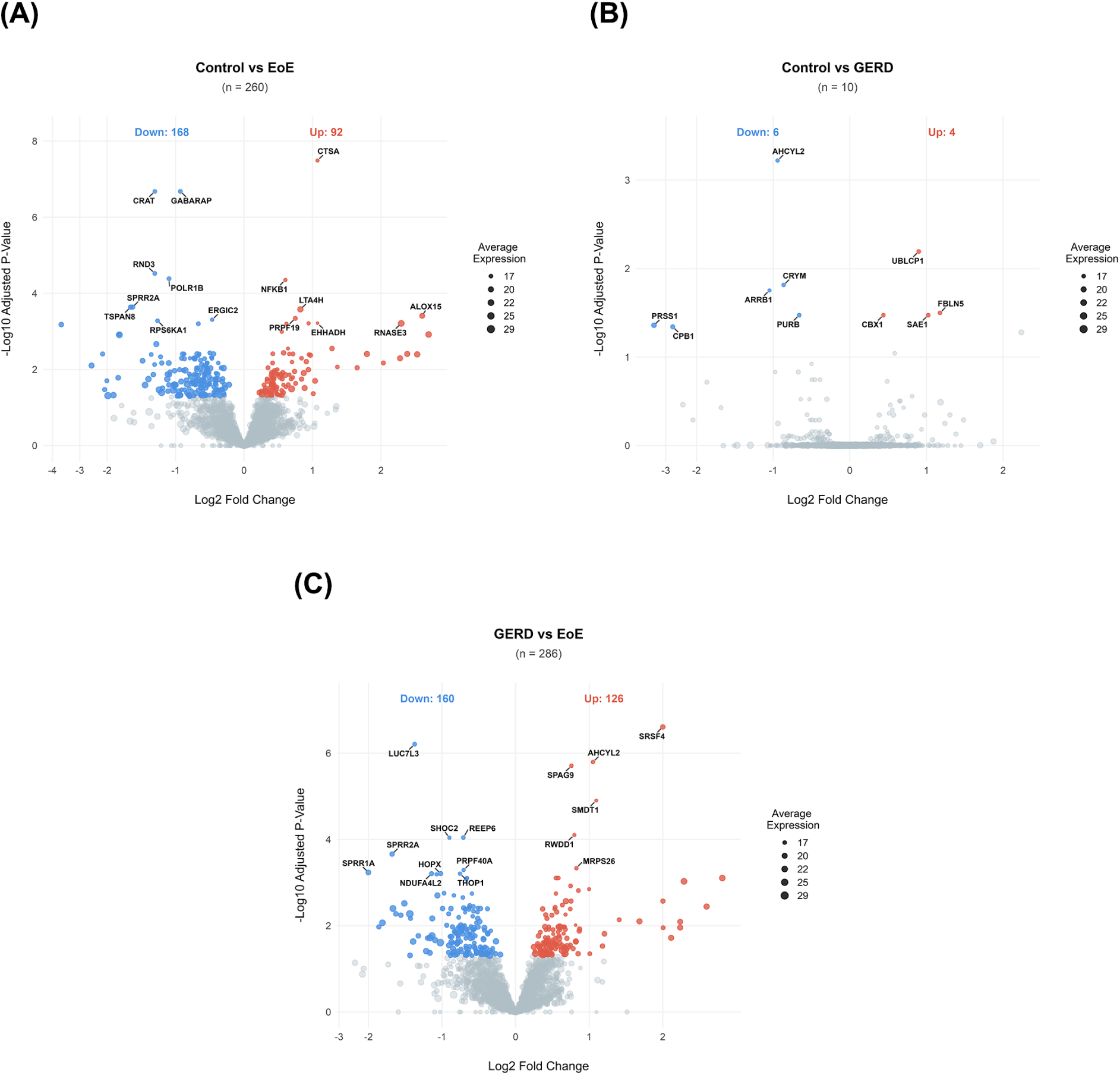
Volcano plots of individual pairwise proteomic comparisons. Volcano plots showing differentially accumulated proteins (DAPs) between (A) C and EoE, (B) C and GERD, and (0) GERD and EoE, using an adjusted P ≤ 0.05. No fold-change threshold was applied to define DAPs in these pairwise comparisons. The gene symbols corresponding to the 15 proteins with the lowest adjusted P-values are highlighted. Dot size represents average protein expression. Abbreviations: C, healthy controls; DAPs, differentially accumulated proteins; EoE, eosinophilic esophagitis; GERD, gastroesophageal reflux disease.

## Notes

### Competing Interest Statement

The authors have declared no competing interest.

### Author Declarations

Comite de Etica de la Investigacion con Medicamentos (CEIm) del Hospital Universitario de La Princesa

